# Descriptive characteristics of continuous oximetry measurement in moderate to severe COVID-19 patients

**DOI:** 10.1101/2021.09.26.21264135

**Authors:** Jonathan A. Sobel, Jeremy Levy, Ronit Almog, Anat Reiner-Benaim, Asaf Miller, Danny Eytan, Joachim A. Behar

## Abstract

Non-invasive oxygen saturation (SpO2) is a central vital sign that supports the management of COVID-19 patients. However, reports on SpO2 characteristics are scarce and none has analysed high resolution continuous SpO2 in COVID-19. We provide the first analysis of high resolution SpO2 across the spectrum of COVID-19 disease severity and respiratory support. A total of 367 COVID-19 patients’ recordings, comprising 27K hours of continuous SpO2 data, could be retrieved from patients hospitalized at the Rambam Health Care Campus. Using oximetry digital biomarkers (OBM), we quantified SpO2 characteristics and showed that the percentage of time under 93% oxygen saturation threshold is the best single OBM discriminating between critical and non-critical patients. OBMs traditionally used in the field of sleep medicine research, were informative for assessing the patient’s response to respiratory support. In addition, periodicity and hypoxic burden biomarkers were affected up to several hours before the initiation of the mechanical ventilation. Characteristics from high resolution SpO2 signal may enable to anticipate clinically relevant events, monitoring of treatment response and may be indicative of future deterioration.x

## Introduction

The ongoing coronavirus disease 2019 (COVID-19) pandemic has spread all over the world and caused, as of early August 2021, over 4,3 million deaths. Approximately 15-20% of confirmed cases develop severe disease, while the fatality rate is 2-5% ^1–3^ depending on the country and the monitored period.

In hospitalized patients with acute respiratory infections, such as patients with COVID-19, continuous SpO2 monitoring has become central and is used to detect desaturation events required for patient triage, risk stratification and escalation of treatment, and to allow the clinicians to track the response to therapeutic interventions such as oxygen enrichment or ventilation ^4,5^. Specifically in COVID-19 patients, SpO2 is often used in the ROX score (ratio of SpO2/FIO2 to respiratory rate) or in the SpO2/FiO2 ratio to predict the future need for oxygen support or mechanical ventilatory support and the success of a therapy ^6–8^. Nevertheless, these studies rely mostly on admission data or discrete measurements during hospitalization. To our knowledge, no study investigated continuous high resolution SpO2 characteristics (patterns and dynamics) in COVID-19 patients.

The present work aims to describe high resolution SpO2 signal characteristics and potential usage along the treatment course for non-critical, critical with or without oxygen support and critical mechanically ventilated patients. The specific objectives of this work are: (1) Determining global SpO2 signal characteristics correlated with disease severity and/or the level of respiratory support; (2) Investigating the optimal definition of a relative or absolute threshold for defining clinically relevant desaturations; (3) Defining characteristics of the SpO2 signal including desaturation parameters and oximetry derived digital biomarkers (OBMs) allowing for the differentiation between critical and non-critical groups or those affected by the level of support; (4) highlighting the potential of OBMs extracted from high resolution SpO2 signal for the detection of early signs of deterioration leading to the indication of mechanical ventilation and for tracking patient’s response to medical treatment.

## Results

### Severe COVID-19 patients and hospitalisation course

Patients were split into two groups based on disease severity with 162 critical and 205 non-critical patients (Table 1). Male patients were prevalent in both groups (60,5% in the non-critical group and 72,8% in the critical group). The in-hospital mortality was 38,9% in the critical group. The age distribution was significantly different (p-value < 0,05) with more patients 65-74 years old in the critical group and less patients 18-44 years old with respect to the non-critical group. The BMI distribution was balanced between groups. The length of stay (Table S1, Figure S3) was significantly longer in the critical group (p-value < 0,001, median non-critical = 5 days, median critical = 14 days). Patients in the critical groups had significantly more overall comorbidities (p-value = 0,01, Table 1, Table S2). Regarding vital signs at admission (Table S1), critical patients depicted a lower room saturation (SpO2 measured breathing room air) and oxygen saturation (SpO2 measured under oxygen support). The respiratory rate was significantly higher in the critical group indicating tachypnea.

**Table 1.** Population sample characteristics. Abbreviation: BMI body mass index

| variable |  | non-critical |  | critical |  | FDR adjusted p-value |
| --- | --- | --- | --- | --- | --- | --- |
|  |  | (n = 205) |  | (n = 162) |  |  |
| gender | Male | 124 | (60,5%) | 118 | (72,8%) | 0,07 |
|  | Female | 81 | (39,5%) | 44 | (27,2%) |  |
| in-hospital mortality | FALSE | 205 | (100%) | 99 | (61,1%) | 6,40e-21 |
|  | TRUE | 0 | (0%) | 63 | (38,9%) |  |
| age group | 18-44 | 28 | (13,7%) | 8 | (4,9%) | 0,02 |
|  | 45-54 | 24 | (11,7%) | 28 | (17,3%) |  |
|  | 55-64 | 51 | (24,9%) | 28 | (17,3%) |  |
|  | 65-74 | 35 | (17,1%) | 44 | (27,2%) |  |
|  | 75+ | 67 | (32,7%) | 54 | (33,3%) |  |
| BMI group | <=20 | 4 | (2,1%) | 2 | (1,3%) | 0,74 |
|  | 20-25 | 42 | (21,6%) | 27 | (17,2%) |  |
|  | 25-30 | 69 | (35,6%) | 57 | (36,3%) |  |
|  | 30< | 79 | (40,7%) | 71 | (45,2%) |  |
| comorbidity | FALSE | 67 | (32,7%) | 29 | (17,9%) | 0,01 |
|  | TRUE | 138 | (67,3%) | 133 | (82,1%) |  |

### Qualitative analysis of SpO2 characteristics

A typical hospital course of a patient after admission is represented in Figure 1A. Hospitalized patients are monitored in the ward or in ICU. Qualitatively, the continuous monitoring of SpO2 of a single patient depicted frequent small desaturations in a non-critical case, while a critical patient presented prolonged events of low SpO2 (Figure 1B). The initiation of mechanical ventilation was visible on the EtCO2 channel. The dynamics of the SpO2 signal was noticeably impacted by mechanical ventilation, with a higher SpO2 level and reduced variability (Figure 1C).

**Figure 1.**
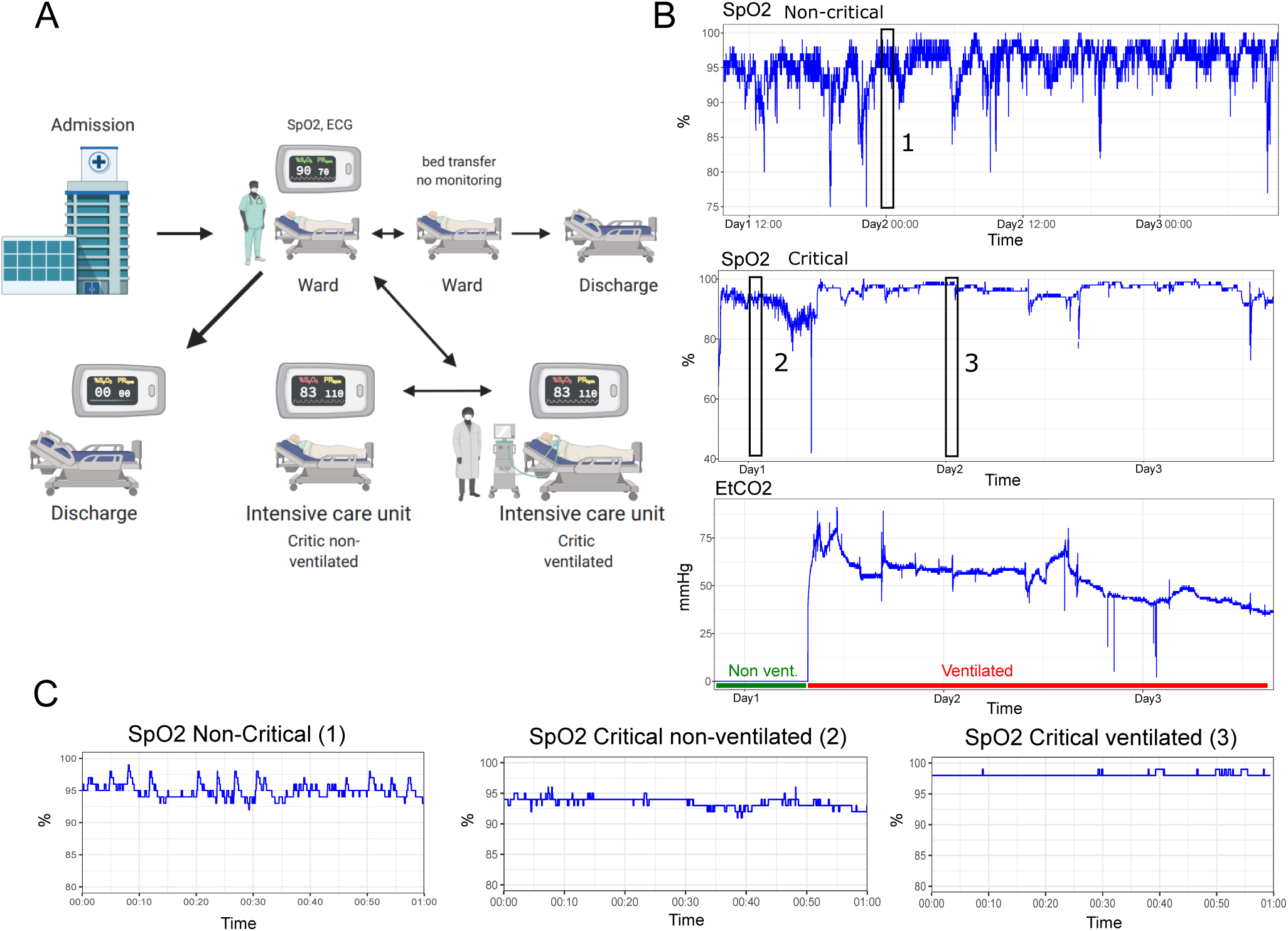
Patient in-hospital movement and examples of SpO2 signal in critical and non-critical cases. A) Frequent in-hospital movement of ward monitored patients. For non-severe patients monitoring is only performed part time and a patient might be transferred or discharged depending on his health status. B) SpO2 and EtCO2 signals of a non-critical and a critical COVID-19 patient with an initiation of ventilation. The transition between non-ventilated and ventilated state is clearly visible after an event of intense desaturation. Non-mechanically ventilated area is indicated in green and mechanically ventilated area is indicated in red. On the EtCO2 channel C) SpO2 signal of the same patients zoomed in on 1h intervals, highlighted by black rectangles in panel B.

### Quantitative analysis of SpO2 characteristics

#### SpO2 distribution

The mean SpO2 was significantly lower among critical patients without support, compared to the non-critical group (p-value < 0,05, 2A,C). The lower SpO2 range between 80% and 90% depicted a higher density in the critical group (p-value < 0,01). The non-critical group showed a narrower peak of densities centered around 96% SpO2. A similar, although milder trend was observed in the critical group versus non-critical patients by oxygen delivery devices (Figure 2B,C). The median oxygen flow rate used for the oxygen support was significantly higher in critical patients (p-value < 0,001). The use of mechanical ventilation reduced the density of low SpO2 (80%-90%) to a level closer to that seen in non-critical patients. On the other hand, critically ill patients who were mechanically ventilated or on oxygen support depicted a higher density of SpO2 level between 97% and 100% compared to critical non ventilated and non-critical patients. The SpO2 density per patient in each group (Figure 2C) revealed the variety of SpO2 patterns, notably in non-critical and critical patients without support. In addition, this analysis revealed that a sub-group of critical patients on oxygen support did not respond well to the therapy and thus subsequently required a mechanical ventilation support.

**Figure 2.**
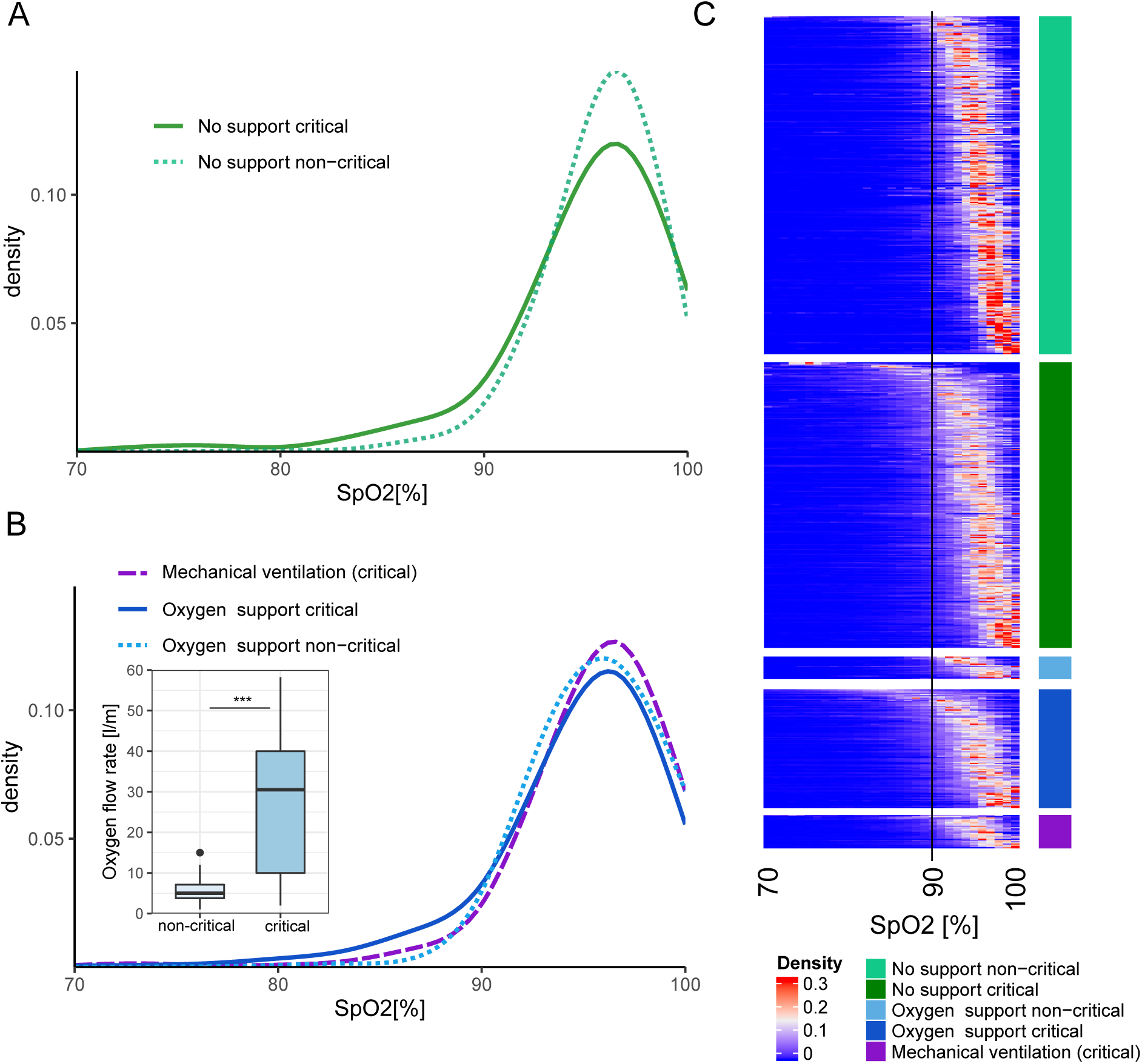
Global characteristics of SpO2 signal. A) Density distribution of SpO2 for non-critical and critical patients without support. B) Density distribution of SpO2 from non-critical and critical patients for intervals under oxygen support or mechanical ventilation. The level of oxygen support (oxygen flow rate) was compared between non-critical and critical patients. The center of boxplot indicates the median, and the bottom and top edges indicate the 25th and 75th percentiles, points indicates outliers. C) SpO2 density heatmap for each interval of SpO2 signal of patient in each group. A vertical black line represent the threshold of 90% SpO2 recommended in the WHO guidelines ^5^ to identify severe patients.

#### Desaturation analysis

Overall, based on the hypoxic burden (Figure 3A), the absolute threshold of 93% was the most discriminating between critical and non-critical groups with p-value < 0,001 and fold-change (FC) of 1,6 for no support and p-value < 0,05 and 2,9 FC under oxygen support. A relative threshold of 3% SpO2 was able to discriminate critical from non-critical patients without support only (p-value < 0,001). Next, for the optimal definition (93% absolute), we compared the distribution of area, depth, and desaturation duration (Figure 6, Figure 3B) between the critical and the non-critical groups and with or without oxygen support. Interestingly, the non-critical group depicted more desaturations per hour compared to the critical groups (p-value < 0,05, FC 1,8). This difference was abolished by oxygen support (non-significant, NS). Similarly the depth and the “desat. time” of the desaturations were significantly more pronounced comparing the critical and non-critical groups when there was no support (p-value < 0,001, FC 1,35). This effect was not significant under oxygen support (NS). The only desaturation parameter that showed a significant difference between those groups under oxygen support was the desaturation area (p-value < 0,05, FC 1,6) consistent with what we observed looking at the hypoxic burden. Overall, the critical group showed a larger depth (p-value < 0,001, 1,23 FC), area (p-value < 0,001, 1,44 FC) and “desat. time” duration (p-value < 0,001, 1,35 FC) of the events with respect to the non-critical group. Oxygen support had a limited effect on the depth and the “desat. time”, no significant differences between oxygen support and no-support were observed. Mechanical ventilation depicted a strong effect by significantly reducing the frequency of desaturations (p-value < 0,001, 1,85 FC) and the depth (p-value < 0,05, 1,21 FC).

**Figure 3.**
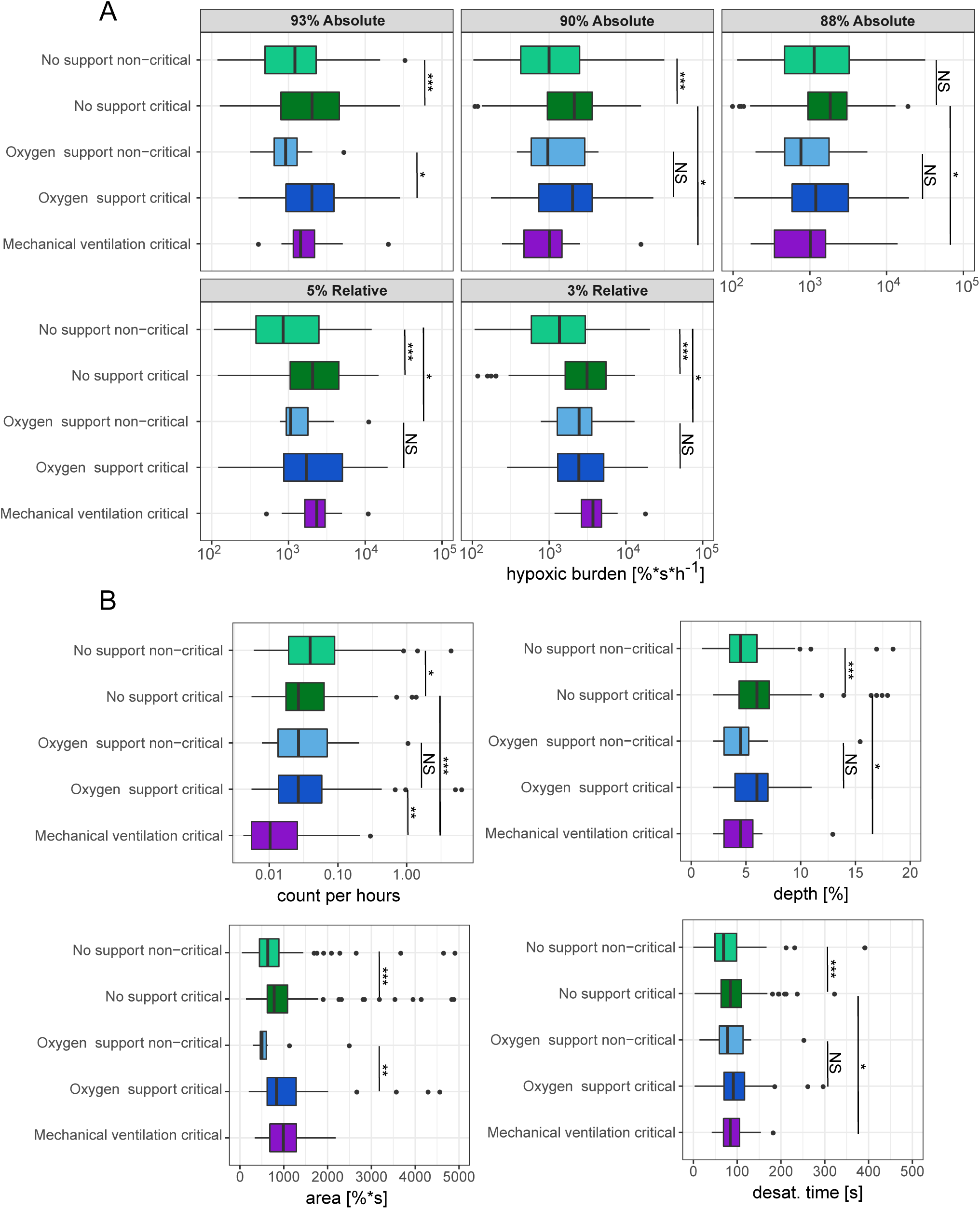
Desaturation definition and parameters A) Hypoxic burden for each relative or absolute desaturation definition. The hypoxic burden was defined as the sum of desaturation areas normalised by recording duration. The hypoxic burden was based on relative or absolute threshold and was measured under oxygen support, mechanical ventilation or without support in critical and non-critical patients. B) Desaturation characteristics for critical and non-critical patients with or without oxygen or mechanical support. The results are shown for an absolute desaturation threshold of 93%. The number of desaturation per hour, the depth the area and the desaturation time (duration between the beginning and the minimum) are represented for each group. *** Wilcoxon test p-value < 0,001,** p-value < 0,01,* p-value < 0,05, NS non-significant. The center of boxplot indicates the median, and the low and high edges indicate the 25th and 75th percentiles, points indicates outliers.

#### Effect of treatment and OBMs

OBMs were investigated using a volcano plot analysis comparing between the critical and non-critical groups without support (Figure 4A). OBMs definitions are presented in Table S3. OBMs related to hypoxic burden and desaturation depicted a large effect (log2 fold change) with highly significant p-values (large log10 p-values) consistent with the preceding results. Remarkably, the integral of SpO2 below the 93% SpO2 (*CA*93) and the percentage of time below the 93% SpO2 (*CT* 93, Figure S6) were the most discriminating OBMs. The same analysis under oxygen support (Figure 4B) revealed that most OBMs related to desaturation and hypoxic burden were not significantly different between the two groups apart from *CT* 93 and *CA*93. Overall, OBMs depicted reduced FDR adjusted p-values under oxygen support. However, general and periodicity related OBMs remained significantly affected between non-critical and critical group. Next, the effect of the support was investigated (Figure S5A-C). In non-critical patients the effect of oxygen support was mild (Figure S5A). Both FC and FDR adjusted p-values ranges were smaller than in the critical versus non-critical comparison (Figure 4A). OBMs related to hypoxic burden and desaturation parameters were still higher under oxygen support (Figure S5A). In the critical group under oxygen versus no support, the picture was similar indicating that the oxygen support had only a mild effect on OBMs (Figure S5B). On the other hand (Figure S5C), mechanical ventilation depicted a more pronounced effect on OBMs, notably with a reduction of all OBMs related to hypoxic burden, desaturation parameters, periodicity and complexity.

**Figure 4.**
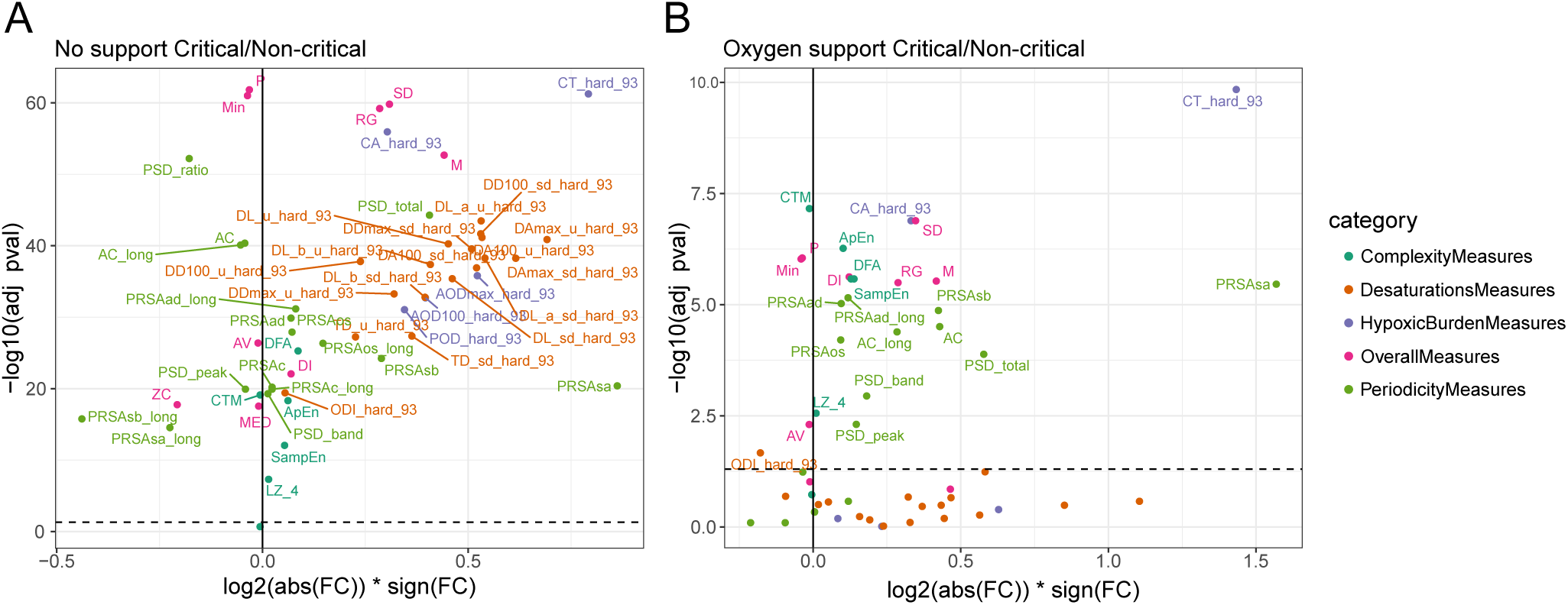
OBMs across the spectrum of disease severity and treatment support. A) Critic versus non-critical without support and B) critical versus non-critical under oxygen support. OBMs definition are available in Table S3. See the Figure S5 for the comparisons between no-support and oxygen support or mechanical ventilation.

### Transition analysis and OBMs’ trajectory

There were a total of 43 patients with transitions, i.e. initiation of mechanical ventilation, with a total of 68 transitions overall. Among the patients that had transitions, they had a Q1 of 1, and a Q3 of 2 transitions, and the maximum number of transitions for a single patient was 3. Panel A of figure S4 shows examples of transition from three representative patients and panel B shows the average profile of every transition detected.

Temporal tracking of selected OBMs is illustrated in figure 5. The median SpO2 depicted a larger variation about 1h before the transition. The desaturation class represented by the *ODI*93, depicted large variations around the transition. The *ODI*93 showed a decreased number about one hour and a half before the transition suggesting larger desaturations in agreement with *CA*93 (hypoxic burden) showing an increase and a high variance before the transition. Approximate entropy *ApEn* (complexity) depicted a small reduction about 4 hours before the transition and marked decreased after the transition. Finally, *PSD*_*total*_ (Periodicity) showed an increasing perturbation between 4 to 1,5 hours before the transition. Thus, the spectral band of interest (0, 014 − 0, 033*Hz*) appeared to have a greater power than the rest of the signal, before the transition.

**Figure 5.**
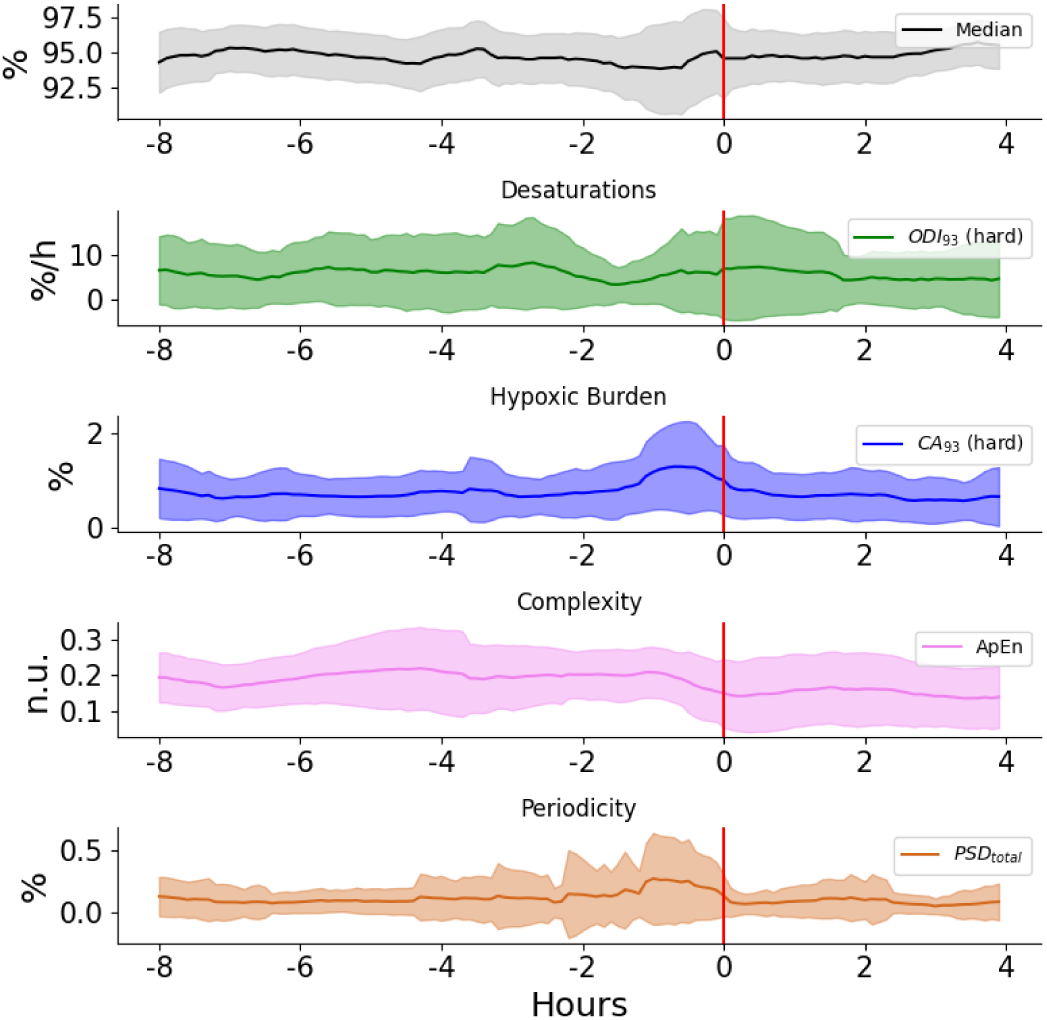
Temporal tracking of OBMs before and after the initiation of mechanical ventilation. OBMs were computed using a sliding windows of 30 minutes, from 8 hours before the transition, to 4 hours after it with a shift of five minutes. The median and IQR were computed over 68 transition events. For each of the five category, one biomarker with low *p value* is represented. OBMs definition are available in Table S3

## Discussion

The oximetry has been widely used for the management of respiratory pathologies, such as pulmonary edema, Acute respiratory distress syndrome (ARDS), pneumonia, obstructive sleep apnea (OSA) ^9–12^, and more recently, in a research setting, for nocturnal chronic obstructive pulmonary disease (COPD) diagnosis ^13^. However, to our knowledge, no study investigated continuous high resolution SpO2 characteristics (patterns and dynamics) in COVID-19 patients.

Patients with COVID 19 presenting to the emergency department go through meticulous investigation (Figure 1A) that includes physical examination, laboratory tests and continuous monitoring of vital signs ^14,15^. Depending on the results of this screening, the patient can be discharged or admitted for observation and treatment in a monitored or non-monitored bed. Oxygen support can be applied in case of hypoxaemia (SpO2 < 94%, or a drop > 3% SpO2) or in dyspnoeic patients with increased work of breathing who are at risk of respiratory failure ^16^. In more severe cases, the patient might be transferred directly to the ICU ^17^. If the patient shows signs of respiratory failure on admission or develops one later, they may be mechanically ventilated, or undergo an attempt of non-invasive support. Thus, multiple decisions are made by physicians in this process who need to factor both subjective observations and objective parameters such as SpO2. However, how these decisions are made and what should be taken into consideration for increasing the level of care is not always clear ^18^. Furthermore, there is a need to sort critical from non-critical patients and identify patients’ status deterioration using non-invasive measurements. Pulse oximetry offers many advantages as it is a cost effective continuous monitoring method that can be used in hospital or in a telemedicine setting ^19,20^.

While healthy people generally depict an oximetry in the range 94% to 98% ^21^, critical COVID-19 patients showed higher density of low SpO2 (80%-90%) as compared to non-critical or critical ventilated patients (Figure 2). Thus several questions arise: what is an appropriate SpO2 threshold to distinguish critical from non-critical patients? what is a good predictor of oxygen support and mechanical ventilation? How does ventilation and oxygen support affect the SpO2 level and its dynamics? Several studies have started to investigate these questions. A recent study showed that a saturation below 90% correlates with a higher probability for mortality ^22^, but this work was based on discrete measures of SpO2 and used death as an endpoint. Another recent investigation of SpO2 and the ROX score at admission concluded that an SpO2 below 78% was a good predictor of mechanical ventilation requirement ^6^ and that a ROX score above 1,4 while on non-invasive ventilation was a good predictor of failure. Other trials suggested that the ratio of SpO2 to FiO2 (or PaO2 to FiO2) could serve as a prognostic marker and could facilitate early adjustment of treatment ^7,23,24^.

In the present study, we motivated that a threshold of 93% best differentiates between critical and non-critical patients under no support or oxygen support. Thus, our work suggests a more stringent threshold compared to the WHO guidelines ^5^ that define an operational threshold of 90% SpO2 to define severe patients. In addition, a drop of 3% SpO2 (relative threshold) was able to discriminate critical from non-critical patients without support. This is consistent with the recommendation of a prompt assessment in the emergency oxygen therapy guidelines ^16^ as it may indicate an acute deterioration in the patient’s condition. In addition, our work highlighted the differences of SpO2 dynamics between the critical and non-critical groups. Specifically, we observed that the non-critical group had frequent shallow desaturations, while critical patients had deeper and longer but less numerous desaturations (Figure 3). Our work also investigated the effect of the oxygen support, which drastically reduced the differences between the two groups. The effect of mechanical ventilation showed a reduction of the SpO2 signal complexity, periodicity, a lower incidence of desaturation and a higher SpO2 saturation level overall with a risk of over oxygenation potentially detrimental ^25,26^.In addition, we have shown that various biomarkers and standard analysis of continuous oximetry formerly developed to study OSA or COPD ^9,13,27,28^ may support the monitoring of COVID-19 patients. Strikingly, two OBMs related to the hypoxic burden class were most discriminative between critical and non-critical patients under no support or under oxygen support. These were: (1) the percentage of time below the 93% oxygen saturation threshold (CT93, Figure S6) and (2) the integral of the signal below the 93% SpO2 level normalized by the total recording time (CA93). This results was consistent with a recent study suggesting the cumulative oxygen deficit as a predictor of mechanical ventilation ^29^.

Our study might have important clinical implications. First, it can serve as a tool to predict the patient’s trajectory while being treated in the ward using oxygen supplementation or non-invasive ventilation. SpO2 monitoring has the advantage of being used frequently and continuously in all patients requiring oxygen treatment, and the data can be saved and processed. Second, assessing oxygenation and desaturation patterns might serve as a prognostic tool in COVID 19 patients. Third, in an overwhelmed medical system, the decision whether to admit or discharge a patient to a ward or ICU is extremely important. Analyzing oxygenation can assist in that manner ^4,12,30^. Finally, our results can be relevant in other medical conditions involving the respiratory system such as pulmonary infections, COPD, ARDS and more. Large scale, preferably prospective randomized trials are required to validate our results.

The presented work has several important limitations. First, our work is a single center retrospective and descriptive study. Second, we did not explore the effect of sub-type of support or stratified parameters such as FiO2, EtCO2, PEEP or pressure support levels.

We provide the first report of continuous SpO2 analysis in COVID-19 patients across severity categories and respiratory support levels. Continuous monitoring of SpO2 is of paramount importance to characterize and manage COVID-19 patients. Mechanical ventilation and oxygen support have an impact on the SpO2 signal characteristics. Finally, OBMs improves the monitoring and enable to anticipate deterioration of the patient’s status.

## Methods

### Study design and participants

This single center retrospective observational cohort study used electronic medical records (EMR) and continuous physiological monitoring data from Rambam Health Care Campus (HCC), a 1000-bed tertiary academic hospital in Northern Israel. During the pandemic the Rambam HCC opened five dedicated COVID-19 departments. The hospital EMR database was queried for hospitalized adult (age 18 and above) cases admitted for COVID-19, from May 1st 2020 until February 1st 2021 with at least one hour of continuous SpO2 recording. During this period, 1810 confirmed COVID-19 cases were admitted to the Rambam HCC (Figure S1). Most cases were mild or moderate and were not necessarily connected to a bedside monitor for continuous surveillance. Five hundred and nineteen patients above 18 years of age were monitored and 367 had more than an hour of SpO2 continuous measurement. Consequently, we collected continuous physiological signal from 162 critical and 205 non-critical COVID-19 patients. Identified waveform data from all COVID-19 units (ward and intensive care unit,ICU) were included. Data were extracted from MINDRAY monitors (Shenzhen, China). Using the MINDRAY software CMSViewer, the available SpO2 data per patient were exported with a resolution of 1Hz. Overall 27K hours of continuous SpO2 signal was collected, including 15K hours on patients breathing room air, 4K hours of mechanically ventilated patients, and 8K hours for patients under oxygen support. Preprocessing of the raw SpO2 signal was performed using a block filter ^28,31^ and a smoothing moving median filter with a window of 9s. EMR data including demographics information for age, sex, weight, body mass index (BMI), length of hospitalization, disease severity, and mortality rates were collected. In addition, monitor information such as the end-tidal CO2 (ETCO2) channel for mechanical ventilation, parameters and timestamps, oxygen support and respiratory information including oxygen flow rate, and respiratory rate were collected. Ethical approval for this research was provided by the local institutional review board under IRB #0141-20.

### COVID-19 definition and severity groups

As in Reiner-Benaim et al. ^32^, and following existing guidelines ^33–35^, COVID-19 positive patients were defined as follow: at least one positive reverse transcription polymerase chain reaction (RT-PCR) test for SARS-CoV-2 in nasopharyngeal swab. Critically ill patients were defined for their entire hospitalization as those who either received mechanical ventilation support (invasive or non-invasive), were hospitalized in an ICU, or were administered vasopressors (Noradrenaline or Vasopressin) or inotropes (Dopamine, Dobutamine, Milrinone, or Adrenaline).

### Oxygen and mechanical ventilatory support

Oxygen support intervals were defined based on the first and the last continuous value of oxygen flow rate extracted from the EMR for an individual patient (Figure S2). Mechanical ventilation intervals were detected using the End-tidal CO2 (EtCO2) channel from the monitors. The EtCO2, which measures the partial pressure of CO2 at end expirium, is recorded only in mechanically ventilated patients. The SpO2 data were split according to an overlap with these predefined intervals. Oxygen delivery and ventilation modes were recorded, including continuous positive airway pressure (CPAP), bi-level positive airway pressure (BIPAP), mask, nasal prongs, T-tube and a tracheostomy mask. Several equipment types were used, notably ventilators such as Hamilton (Hamilton Medical, Bonaduz, Switzerland), ServoAIR, Servo I (Getinge, Gothenburg, Sweden), EVITA (Dräger,Lübeck, Germany), VELA (VYAIRE MEDICAL INC., Chicago, Illinois, United States) and high flow oxygen delivery devices such as Airvo2 (Fisher and Paykel, Auckland, New Zealand) and Vapotherm (Vapotherm Inc., Exeter, New Hampshire, United States).

### Events definition

#### Desaturation events

Two categories of desaturations can be defined: (1) “relative” desaturations corresponding to a decrease of x% (here taken as 3% or 5%) of the SpO2 signal, before returning to 1% below the initial saturation level. The relative threshold desaturation detector is based on the Oxygen Desaturation Index (ODI) which is traditionally used in sleep medicine. The ODI algorithm from Jung et al. ^36^ was used with its implementation and validation by Behar et al. ^37^; (2) “hard” desaturations defined as SpO2 level falling below a given threshold of x% SpO2 (here taken as 93%, 90% or 88%). When the SpO2 signal falls below this value, a desaturation is detected. Figure 6 presents examples of desaturations detected using the hard threshold and relative threshold. The event has to be of a minimum length of 120 seconds. This is to be considered a desaturation event to avoid a false positive reading caused by the patient’s movements or noise. For each detected desaturation, several desaturation parameters were extracted. Specifically area, length, depth, time between the beginning and the minimum point (defined as desat. time), the time between the minimum and the end (defined as resat. time) and the duration between two consecutive desaturations (interval time).

**Figure 6.**
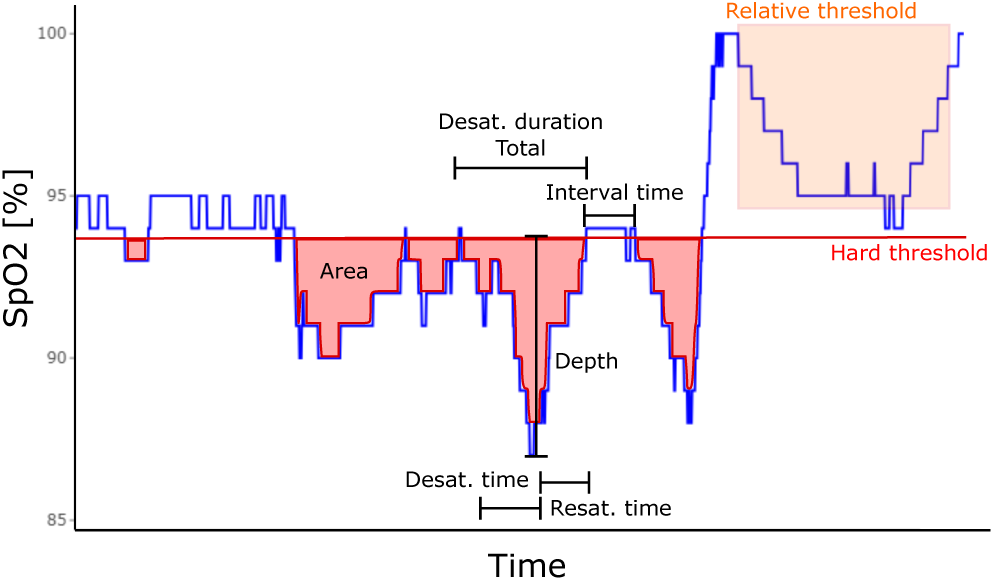
Example of SpO2 signal with desaturation parameters highlighted. Here the hard threshold is at 93% and the relative threshold is of 5%.

#### Transition to mechanical ventilation event

For each patient in the critical group, the time of initiation of mechanical ventilation was extracted on the EtCO2 channel using a sliding window procedure. Briefly, two consecutive windows of five seconds long were created. When sliding the two consecutive windows over the CO2 signal, if the first one had no signal within it whereas the second one had only valid values within it, this was labeled as an event of transition to mechanical ventilation.

#### Oximetry biomarkers (OBMs)

OBMs were extracted from the SpO2 signal with a sampling frequency of 1Hz. As previously described by Levy et al. ^27^, OBMs definitions are presented in Table S3. these biomarkers are divided into 5 categories: (1) General Statistics: time-based statistics describing the SpO2 data distribution; (2) Complexity: quantifies the presence of long-range correlations in non-stationary signal; (3) Periodicity: quantifies consecutive events to identify periodicity in the SpO2 signal; (4) Desaturations: time-based descriptive measures of the desaturation patterns occurring throughout the signal; (5) Hypoxic Burden: time-based measures quantifying the overall degree of hypoxemia imposed on the heart and other organs during the recording period.

### Statistical Analysis

#### Cohort

Comorbidities and demographics including age, weight, BMI, length of stay and death were collected. Comorbidities were defined by ICD-9 codes and analysed as by Benaim et al ^32^. A thorough analysis of comorbidities, demographics and mortality rate was performed to characterize predispositions for both the critical and non-critical group. Demographic variables and comorbidity rates, were compared between severity groups using the Chi-squared test or Fisher’s exact test for categorical variables, and analysis of variance or Kruskal-Wallis test for continuous variables. The p-values across all tests were corrected to control the false discovery rate (FDR) criterion ^38^. Medians and inter-quartile range (IQR) were used to describe the continuous variables.

#### SpO2 global characteristics

The SpO2 signal was summarised using a density of SpO2 for each group of patients investigated. In addition, the SpO2 density was computed per patient and support interval and was represented in a heatmap sorted by SpO2 mean for each group.

#### Desaturation characteristics

In order to decide which desaturation definition was the most clinically relevant, we studied the hypoxic burden as a function of each threshold (Figure 3A). The hypoxic burden was defined by the sum of areas of desaturations per hours for a given patient. The characteristics of the desaturation between each group were compared using the Wilcoxon test.

#### OBMs comparisons across severity and support level

The OBM toolbox ^27^ was used to extract OBMs from the SpO2 signal using consecutive and non-overlapping windows of 1h. The average of each OBM per patient and under each support was computed for analysis. OBMs were used to discriminate between critical and non critical patients and characterize the effects of mechanical ventilation and oxygen support on the SpO2 signal. The goal of this last analysis was to determine the influence of oxygen support or mechanical ventilation on the SpO2 signal. Groups OBMs were compared using using the Wilcoxon test.

#### OBMs and transition events

A sliding window of 30 minutes with a shift of 5 min was applied on the SpO2 signal 8h before to 4h after transitions to study the dynamic of the signal and identify candidate OBMs anticipating a deterioration of the patient requiring intubation and mechanical ventilation. The median and the interquartile range of all transitions are represented for selected OBMs that depicted large variations before the transition.

The R software ^39^ was used for statistical analysis. A significance threshold at 0,05 was used.

## Data Availability

The anonymised database including the waveforms and the clinical data from the COVID-19 patients will be accessible upon reasonable request, which will be individually reviewed by the ethical committee of the Rambam HCC.

## Contributors

J.S., J.B., R.A., A.R.B., and D.E. were involved in the conception and design of the study. J.S: was the coordinator of the study. J.S., R.A., A.M. and A.R.B were responsible for the data collection. J.S. and J.B. wrote the first draft. J.S., J.L. and A.R.B. were in charge of the analysis. A.R.B and R.A. accessed and verified the data. All authors were involved in the interpretation, critically reviewed the first draft, and approved the final version.

## Declaration of interests

The authors declare that there is no conflict of interest.

## Code availability

The source code used in this research is available at physiozoo.com.

## ACKNOWLEDGEMENTS

This research is partially supported by The Milner Foundation, founded by Yuri Milner and his wife Julia. We are grateful to the Placide Nicod fundation for their financial support (J.S.). We acknowledge the financial support of the Technion Machine Learning and Intelligent Systems center (MLIS). We would like to thank Prof. Pierre Singer for insightful discussion regarding the present work. We are grateful to the care team from the COVID-19 units of Rambam HCC.

## Supplementary Information

**Table S1.**
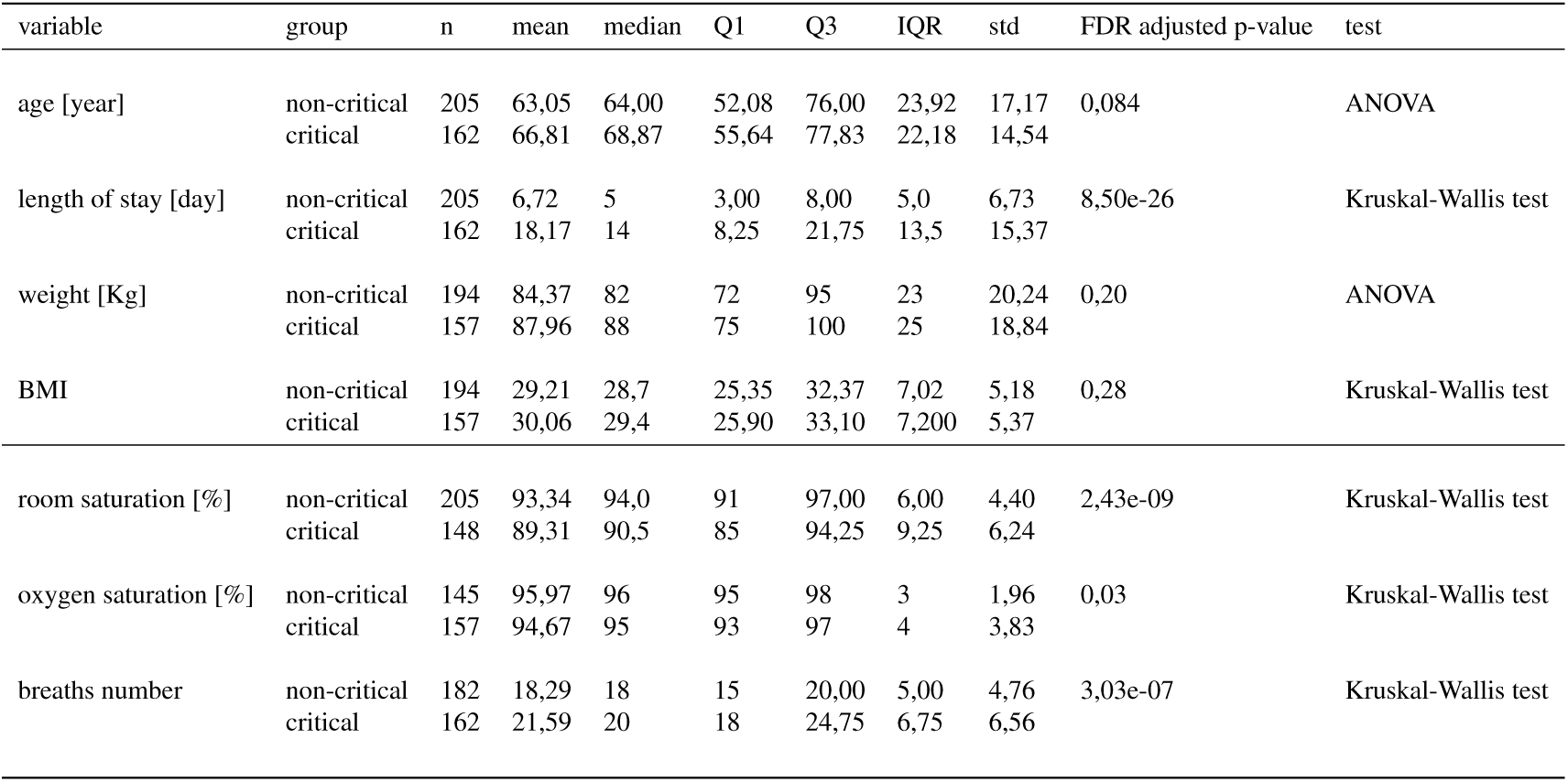
Critical and non-critical patient characteristics: demographic, length of stay, and respiratory parameters at admission.

| variable | group | n | mean | median | Q1 | Q3 | IQR | std | FDR adjusted p-value | test |
| --- | --- | --- | --- | --- | --- | --- | --- | --- | --- | --- |
| age [year] | non-critical | 205 | 63,05 | 64,00 | 52,08 | 76,00 | 23,92 | 17,17 | 0,084 | ANOVA |
|  | critical | 162 | 66,81 | 68,87 | 55,64 | 77,83 | 22,18 | 14,54 |  |  |
| length of stay [day] | non-critical | 205 | 6,72 | 5 | 3,00 | 8,00 | 5,0 | 6,73 | 8,50e-26 | Kruskal-Wallis test |
|  | critical | 162 | 18,17 | 14 | 8,25 | 21,75 | 13,5 | 15,37 |  |  |
| weight [Kg] | non-critical | 194 | 84,37 | 82 | 72 | 95 | 23 | 20,24 | 0,20 | ANOVA |
|  | critical | 157 | 87,96 | 88 | 75 | 100 | 25 | 18,84 |  |  |
| BMI | non-critical | 194 | 29,21 | 28,7 | 25,35 | 32,37 | 7,02 | 5,18 | 0,28 | Kruskal-Wallis test |
|  | critical | 157 | 30,06 | 29,4 | 25,90 | 33,10 | 7,200 | 5,37 |  |  |
| room saturation [%] | non-critical | 205 | 93,34 | 94,0 | 91 | 97,00 | 6,00 | 4,40 | 2,43e-09 | Kruskal-Wallis test |
|  | critical | 148 | 89,31 | 90,5 | 85 | 94,25 | 9,25 | 6,24 |  |  |
| oxygen saturation [%] | non-critical | 145 | 95,97 | 96 | 95 | 98 | 3 | 1,96 | 0,03 | Kruskal-Wallis test |
|  | critical | 157 | 94,67 | 95 | 93 | 97 | 4 | 3,83 |  |  |
| breaths number | non-critical | 182 | 18,29 | 18 | 15 | 20,00 | 5,00 | 4,76 | 3,03e-07 | Kruskal-Wallis test |
|  | critical | 162 | 21,59 | 20 | 18 | 24,75 | 6,75 | 6,56 |  |  |

**Table S2.** Comorbidities prevalence among study patients at admission and comparison between critical and non-critical groups.

| variable |  | non-critical |  | critical |  | FDR adjusted p-value |
| --- | --- | --- | --- | --- | --- | --- |
|  |  | (n = 205) |  | (n = 162) |  |  |
| cancer ALL | FALSE | 190 | (92,7%) | 145 | (89,5%) | 0,48 |
|  | TRUE | 15 | (7,3%) | 17 | (10,5%) |  |
| cancer SOLID | FALSE | 191 | (93,2%) | 146 | (90,1%) | 0,45 |
|  | TRUE | 14 | (6,8%) | 16 | (9,9%) |  |
| cancer HEMATOLOGIC | FALSE | 204 | (99,5%) | 160 | (98,8%) | 0,69 |
|  | TRUE | 1 | (0,5%) | 2 | (1,2%) |  |
| acute renal failure | FALSE | 199 | (97,1%) | 153 | (94,4%) | 0,40 |
|  | TRUE | 6 | (2,9%) | 9 | (5,6%) |  |
| CKD | FALSE | 183 | (89,3%) | 132 | (81,5%) | 0,14 |
|  | TRUE | 22 | (10,7%) | 30 | (18,5%) |  |
| asthma and bronchiectasis | FALSE | 197 | (96,1%) | 158 | (97,5%) | 0,68 |
|  | TRUE | 8 | (3,9%) | 4 | (2,5%) |  |
| COPD | FALSE | 195 | (95,1%) | 153 | (94,4%) | 0,81 |
|  | TRUE | 10 | (4,9%) | 9 | (5,6%) |  |
| dementia | FALSE | 201 | (98%) | 155 | (95,7%) | 0,36 |
|  | TRUE | 4 | (2%) | 7 | (4,3%) |  |
| diabetes | FALSE | 137 | (66,8%) | 93 | (57,4%) | 0,20 |
|  | TRUE | 68 | (33,2%) | 69 | (42,6%) |  |
| hypertension | FALSE | 102 | (49,8%) | 76 | (46,9%) | 0,74 |
|  | TRUE | 103 | (50,2%) | 86 | (53,1%) |  |
| hyperlipidemia | FALSE | 133 | (64,9%) | 85 | (52,5%) | 0,07 |
|  | TRUE | 72 | (35,1%) | 77 | (47,5%) |  |
| ischemic heart disease | FALSE | 178 | (86,8%) | 138 | (85,2%) | 0,78 |
|  | TRUE | 27 | (13,2%) | 24 | (14,8%) |  |
| cardiovascular disease | FALSE | 83 | (40,5%) | 52 | (32,1%) | 0,22 |
|  | TRUE | 122 | (59,5%) | 110 | (67,9%) |  |
| smoking | FALSE | 181 | (88,3%) | 140 | (86,4%) | 0,75 |
|  | TRUE | 24 | (11,7%) | 22 | (13,6%) |  |
| stroke | FALSE | 189 | (92,2%) | 143 | (88,3%) | 0,36 |
|  | TRUE | 16 | (7,8%) | 19 | (11,7%) |  |

**Table S3.**
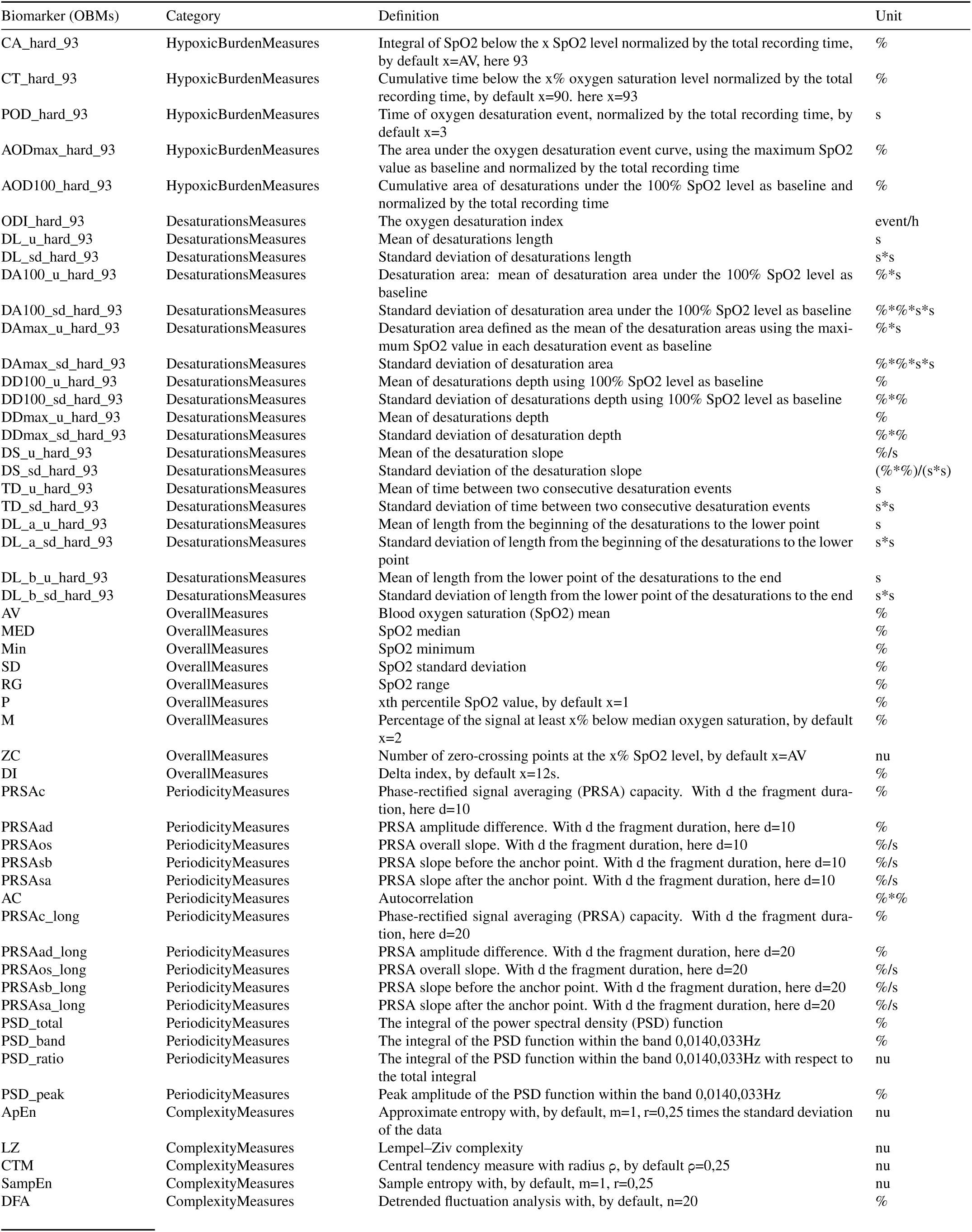
Oximetry derived biomarkers (OBMs) definitions adapted from levy et al. ^27^

**Figure S1.**
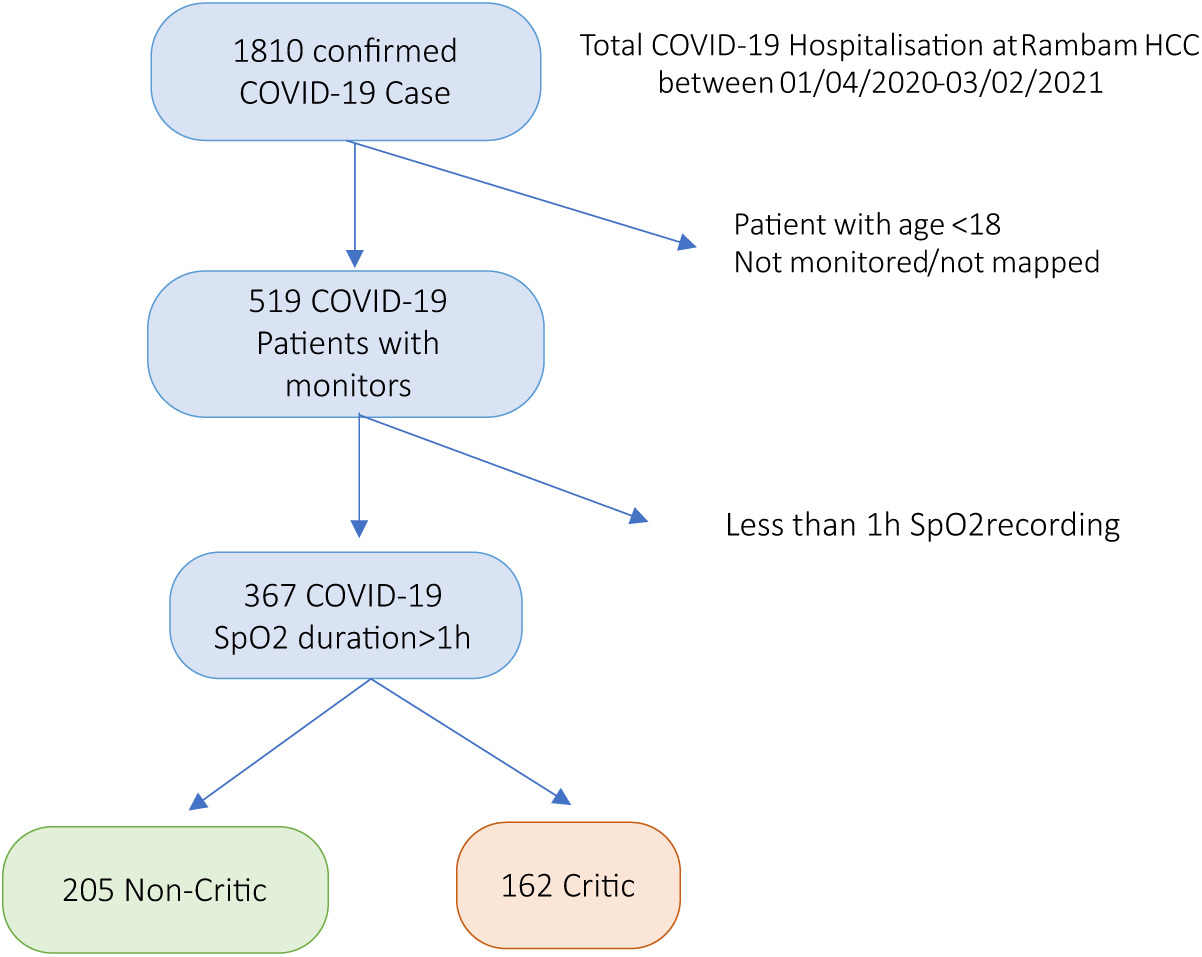
Database content and selection criteria

**Figure S2.**
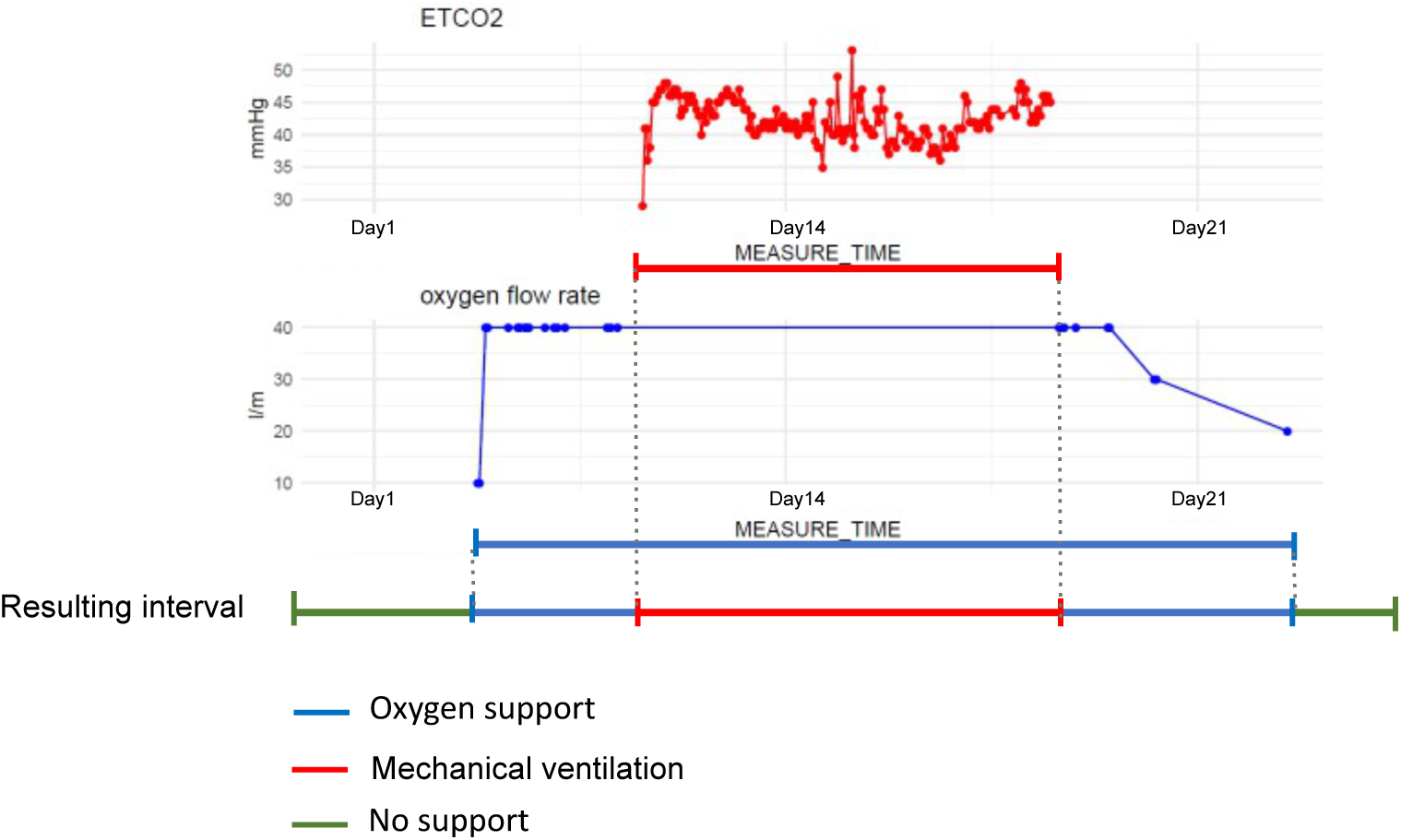
Definition of oxygen support and mechanical ventilation intervals. The first and the last time stamp are used from the oxygen flow rate and from the EtCO2 channel. No support is defined between the admission and the first information about support (or the last and the discharge) or if there is no support information, the whole monitoring is considered without support.

**Figure S3.**
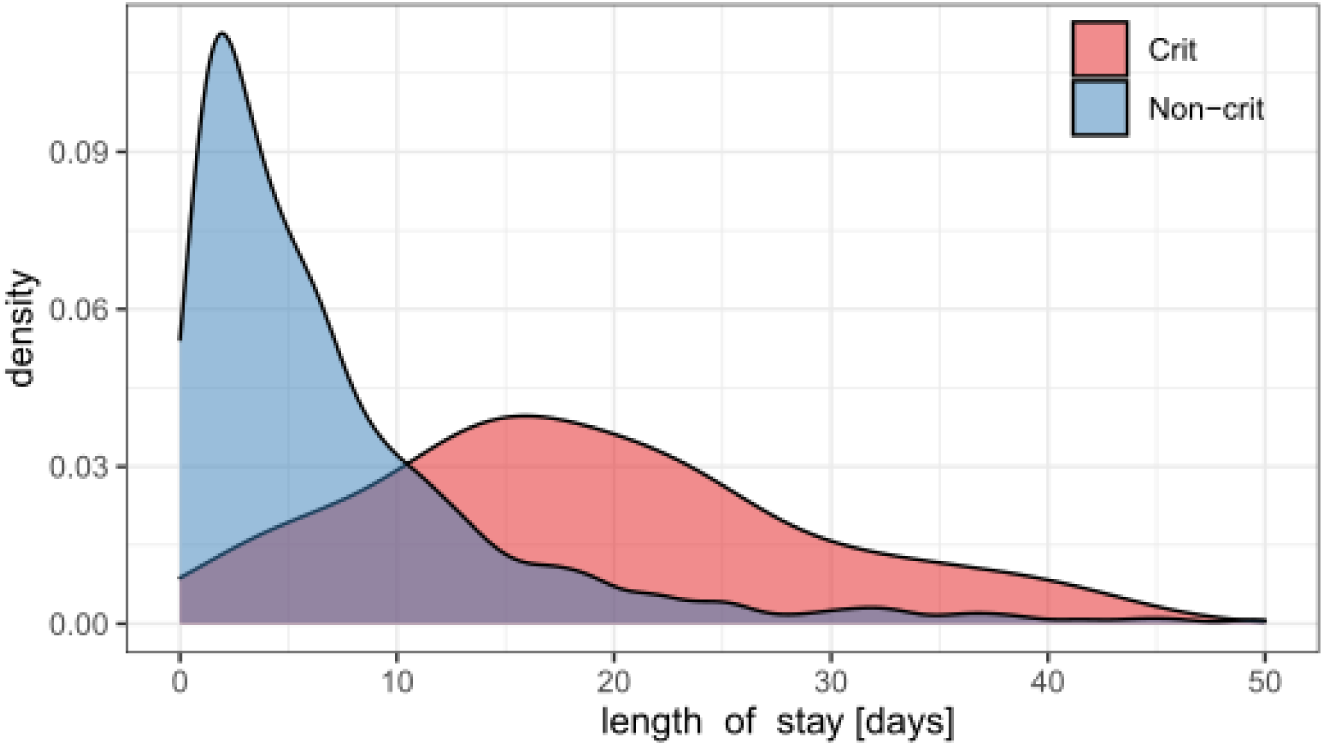
Distribution of length of stay for critical (red) and non-critical (blue) COVID-19 patients.

**Figure S4.**
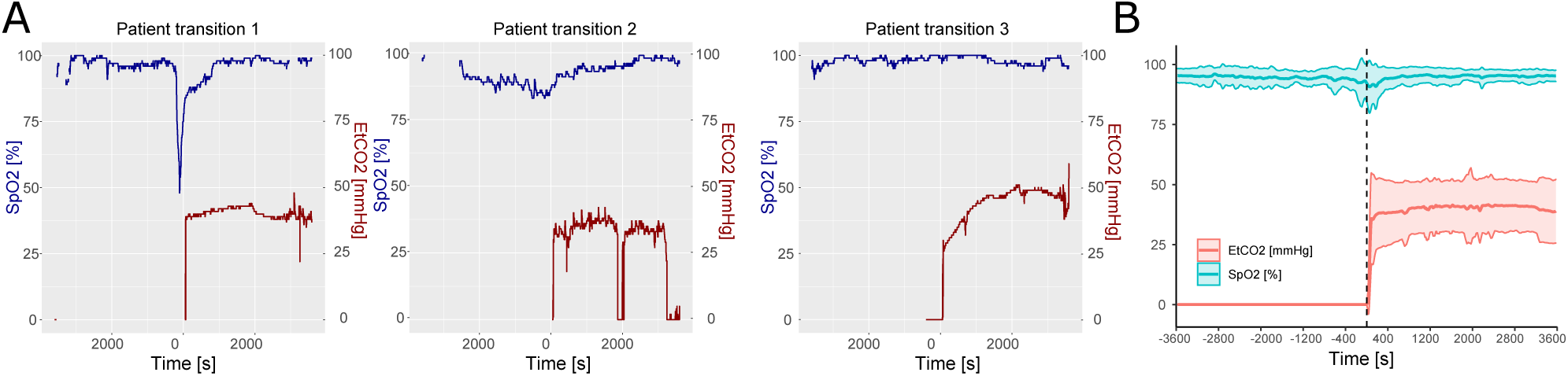
Effect of the ventilation on SpO2 signal at the initiation of the ventilation. A) Representative initiation of ventilation in three different patients. EtCO2 (red) and SpO2 (blue) signal were extracted using 2h window centered on the ventilation initiation. B) Average and standard deviation of EtCO2 and SpO2 for all the detected transitions.

**Figure S5.**
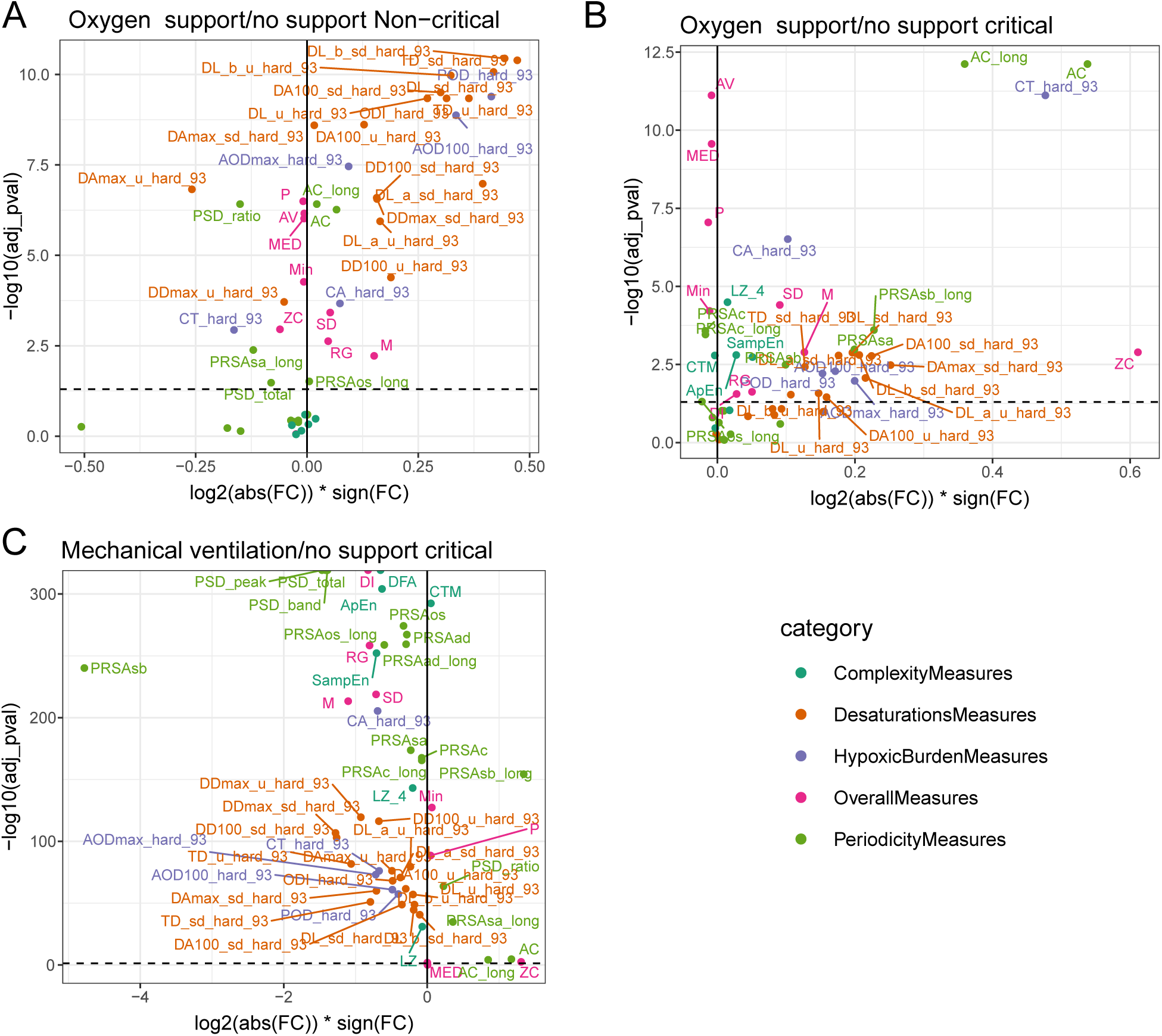
OBMs across the spectrum of disease severity and treatment support. A) Non-critical under oxygen compared to non-critical without support. B) Critic under oxygen compared to critical without support. C) Critic under mechanical ventilation compared to critical without support. OBMs definition are available in Table S3

**Figure S6.**
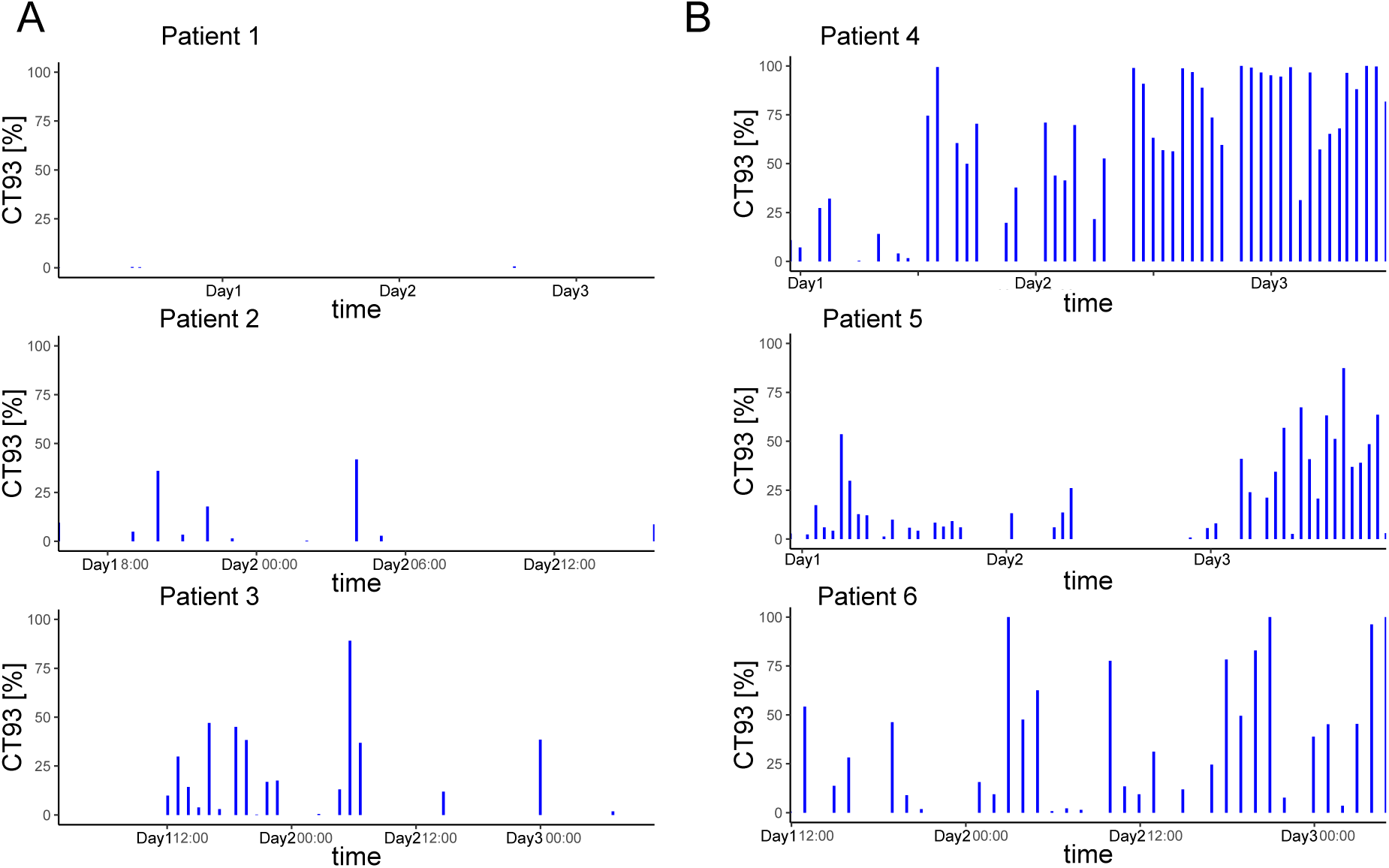
OBM CT93, defined as the percentage of time under the 93% SpO2 threshold, extracted from 1h windows for representative A) non-critical and B) critical patients without support. CT93 was the most discriminating biomarker between critical and non-critical group.

## Notes

### Competing Interest Statement

The authors have declared no competing interest.

### Author Declarations

Ethical approval for this research was provided by the Rambam HCC institutional review board under IRB #0141-20.

### Summary of Updates

Revised abstract and reformatting

